# Improving Deceased Donor Kidney Utilization: Predicting Risk of Nonuse with Interpretable Models

**DOI:** 10.1101/2024.09.11.24313488

**Authors:** Ruoting Li, Sait Tunc, Osman Ozaltin, Matthew J. Ellis

**Affiliations:** Department of Critical Care Medicine, University of Pittsburgh Medical Center, Pittsburgh, PA 15213; Grado Department of Industrial and Systems Engineering, Virginia Tech, Blacksburg, VA 24060; Edward P. Fitts Department of Industrial and Systems Engineering, North Carolina State University, Raleigh, NC 27695; Division of Nephrology, Department of Medicine, Duke University School of Medicine, Durham, NC 27710

**Keywords:** Deceased donor kidney, kidney transplantation, nonuse risk prediction, predictive modeling, clinical decision-making

## Abstract

**Background:** Despite the increasing disparity between the number of patients awaiting kidney transplants and the availability of deceased donor kidneys, a significant number of donated kidneys go unused. Early identification of organs at high risk of nonuse can facilitate effective allocation interventions, ensuring these organs are offered to patients who could potentially benefit from them. While several machine learning models have been developed to predict nonuse risk, the complexity of these models compromises their practical implementation.

**Methods:** We propose implementable nonuse risk prediction models that consist of a minimal set of variables, including the Kidney Donor Risk Index (KDRI), along with factors selected by machine learning models or transplantation experts. Our approach takes into account the influence of Organ Procurement Organization (OPO) behavior on kidney disposition.

**Results:** The proposed models demonstrate competitive performance compared to more complex models that involve a large number of variables. Importantly, they maintain simplicity and interpretability.

**Conclusions:** Our results provide accurate risk predictions, offer valuable insights into key factors contributing to kidney nonuse, and underscore significant variations among OPOs in the allocation of hard-to-place kidneys. These findings can inform the design of effective organ allocation interventions, increasing the likelihood of transplantation for hard-to-place kidneys.

## 1 Introduction

Kidney transplantation is the gold standard treatment for patients with end-stage renal disease. Nearly 80% of kidneys are recovered from deceased donors, however a significant challenge remains: almost 90,000 U.S. patients stay on the waiting list, while *one out of every four* donated kidneys that are recovered for transplantation go unused.^1–3^ Perceived organ quality plays a crucial role in the alarming nonuse rate, as does the intricacies of appropriately matching available organs with suitable recipients.^4^ To alleviate such significant discrepancies, mechanisms to expeditiously match donated organs at higher risk of nonuse with patients who may potentially benefit from receiving them emerge as a pressing need.^5,6^

In current practice, to increase the utilization and expedite the placement of “hard-to-place” kidneys, organ procurement organizations (OPOs) can deviate from the match-run process and extend out-of-sequence offers to transplant centers. The prevalence of such offers has recently increased because latest updates in the kidney allocation system inadvertently created delays in local kidney placements.^7^ Without defined guidelines, allocation exceptions may amplify the existing inequalities in organ access.^8,9^ Thus, predicting which kidneys require additional effort or interventions for successful placement is crucial for making equitable and transparent allocation decisions.

The Kidney Accelerated Placement (KAP) initiative, launched in July 2019, aimed at identifying hard-to-place kidneys and channeling them to transplant centers with a history of accepting such organs.^3,10^ However, the KAP project failed to increase organ utilization due to: (i) a vague definition of “hard-to-place” kidneys, and (ii) delayed acceleration of placement for such kidneys until they had been rejected by multiple local and regional transplant programs.^11^ Our study addresses these two shortcomings by proposing machine learning models that can accurately identify kidneys at high risk of nonuse either *before the beginning* of the match run process or during its *early stages*, enabling *timely interventions*.

The Kidney Donor Risk Index (KDRI) and Kidney Donor Profile Index (KDPI) serve as mainstays for clinicians and transplant decision-makers for evaluating kidney quality and predicting post-transplant longevity, both of which subsequently impact the likelihood of offer acceptance.^11,12^ KDRI is a relative graft failure risk metric, and KDPI maps KDRI to a cumulative percentage scale. These indices have been used by transplant policymakers to assess the nonuse risk of deceased donor kidneys when designing interventions to mitigate the alarming rates of organ nonuse.^13,14^ The KAP project, for example, leveraged minimum 80% KDPI (slightly deviating from the conventional 85% threshold) as a primary criterion for triggering accelerated placement interventions.^13^ However, using KDPI alone for predicting nonuse risk is fallible since KDPI was not designed for this purpose. As illustrated in Figure 1, many kidneys with high KDPI find recipients, while a significant portion with KDPI<85% go unused.

**Figure 1.**
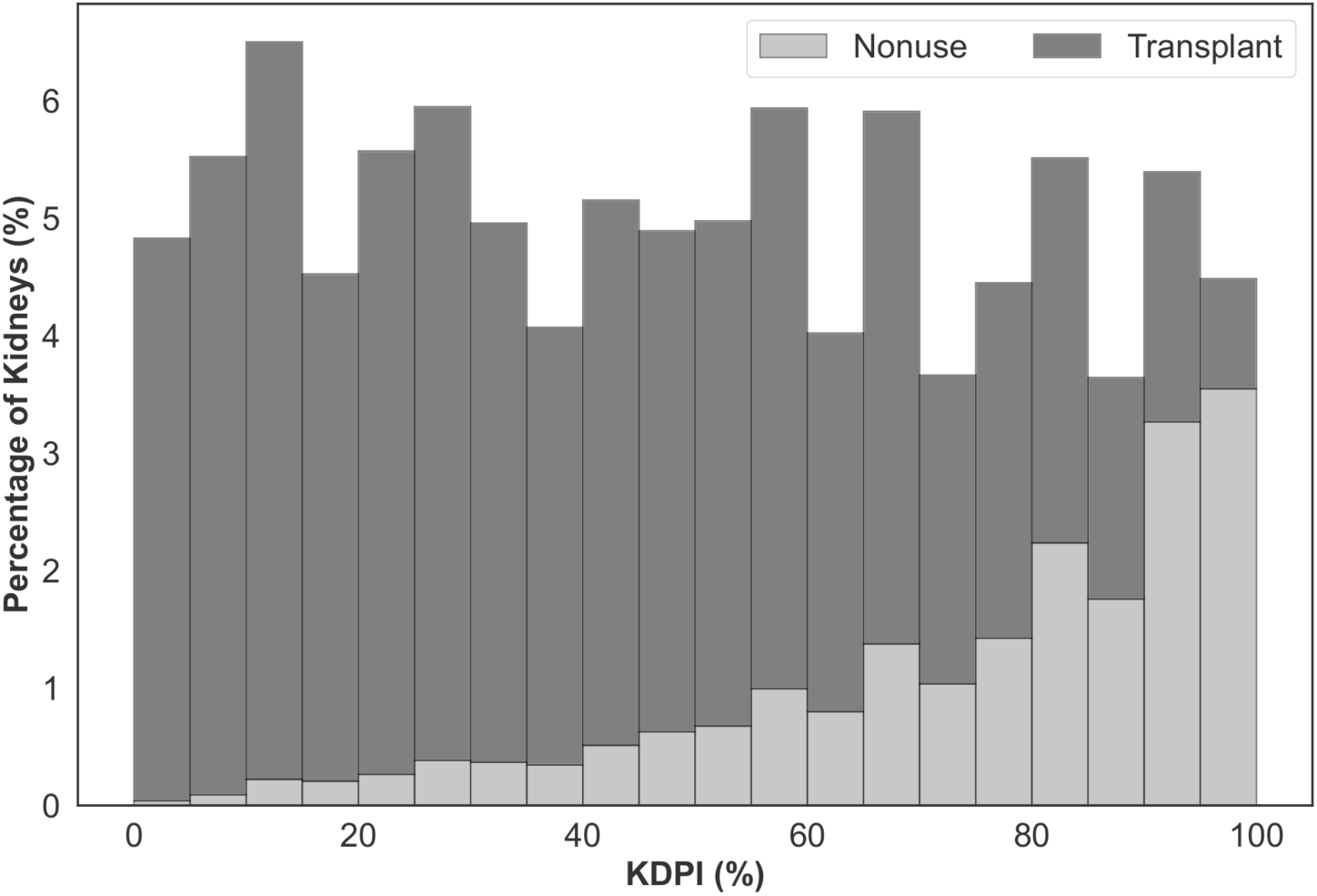
Percentage of deceased donor kidneys recovered for transplantation between 2016 and 2021 that are not utilized (light gray) or transplanted (dark gray) with respect to KDPI.

There exists research advocating for nonuse risk modeling to supplant the KDPI 85% benchmark.^15^ Massie et al.^16^ and Marrero et al.^17^ developed logistic regression models that outperform KDRI in nonuse risk prediction. Barah & Mehrotra^18^ proposed machine learning models, such as random forest, boosting tree, and neural networks. These models demonstrated good predictive performance; however they had three major drawbacks. First, they overlooked the potential benefits of including KDRI/KDPI as a variable. Although these metrics, in isolation, may not provide a reliable nonuse risk assessment, their widespread acceptance and usage in clinical settings cannot be dismissed. Second, they used a multitude of variables, impeding their interpretability and implementability.^19,20^ Finally, they focused on donor and organ characteristics, ignoring the impact of the OPO characteristics on the utilization likelihood of kidneys.^21,22^

We propose implementable and parsimonious machine learning models to predict the nonuse risk of kidneys during the match run. Harmonizing KDRI with a minimal number of variables, the proposed models can help improve kidney utilization rates by informing expedited placement interventions in a timely manner. Our computational experiments demonstrate the competitive performance of the proposed models and identify donor and OPO-level factors affecting the utilization of hard-to-place organs.

## 2 Materials and Methods

### 2.1 Data

Our data set, provided by the United Network for Organ Sharing, includes records from 61,320 deceased donors, between January 2016 and September 2021, who had at least one kidney recovered for transplantation. Each record contains 530 variables, encompassing donor demographic characteristics, physical properties, and relevant medical information, such as laboratory values and comorbidities. Furthermore, we obtained Potential Transplant Recipient (PTR) data for the same time period, which captures all kidney offers made to patients on the US waiting list. The PTR data logs the match run creation time for each donor, which was used to determine ischemia time. We removed 110 donors missing a match run creation time from the analysis.

We identified an initial list of variables linked to kidney nonuse in the literature.^16–18^ This list was further augmented by kidney transplant experts on our team, leading to the list of 36 variables included in our analysis (Table 1). These variables were used to generate an observation vector for each kidney, where each kidney from the same donor was treated as a separate observation (Figure 2).

**Figure 2.**
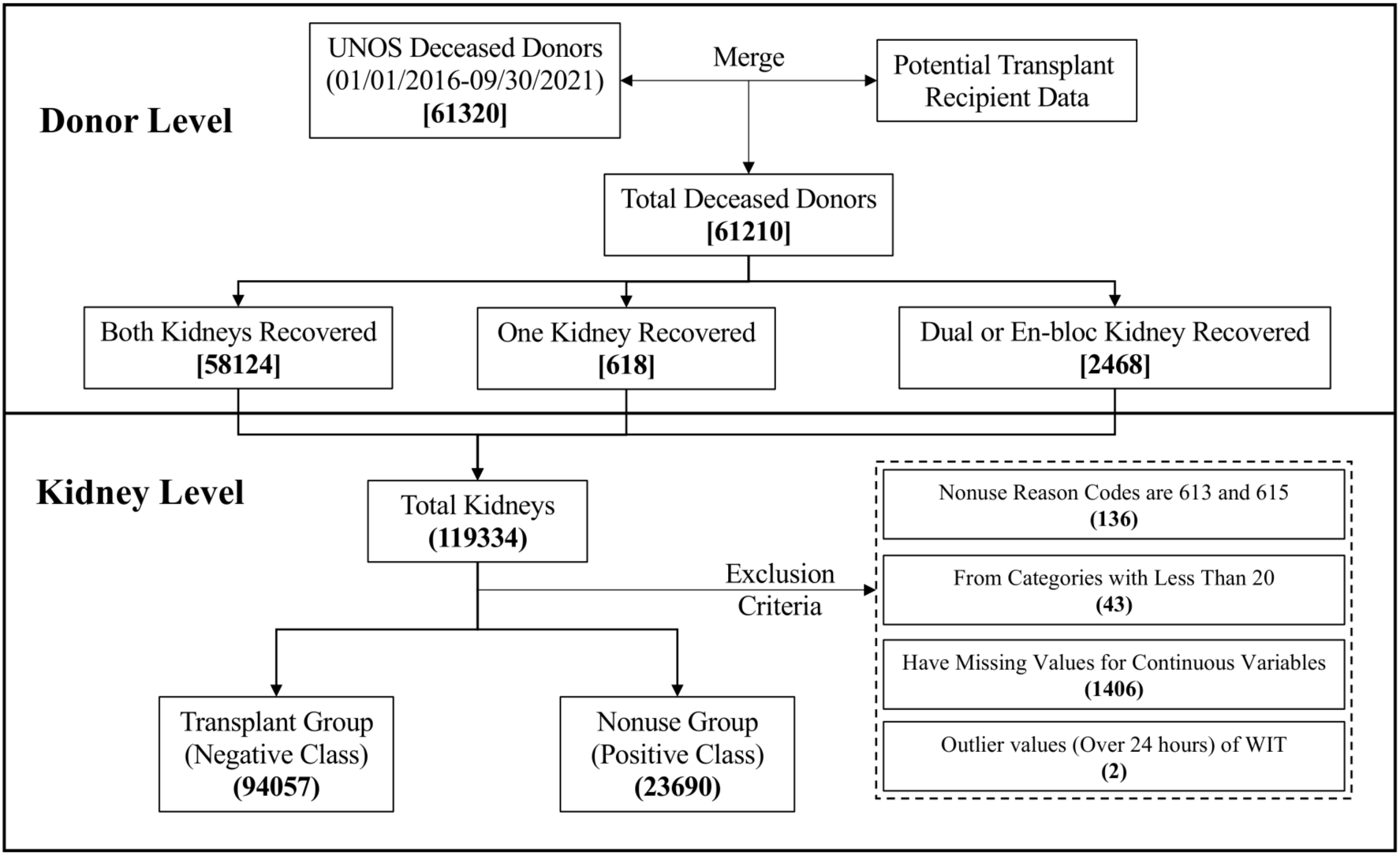
The process for creating the kidney-level data set. Numbers inside the square brackets and parentheses indicate the number of donors and kidneys, respectively.

**Table 1:**
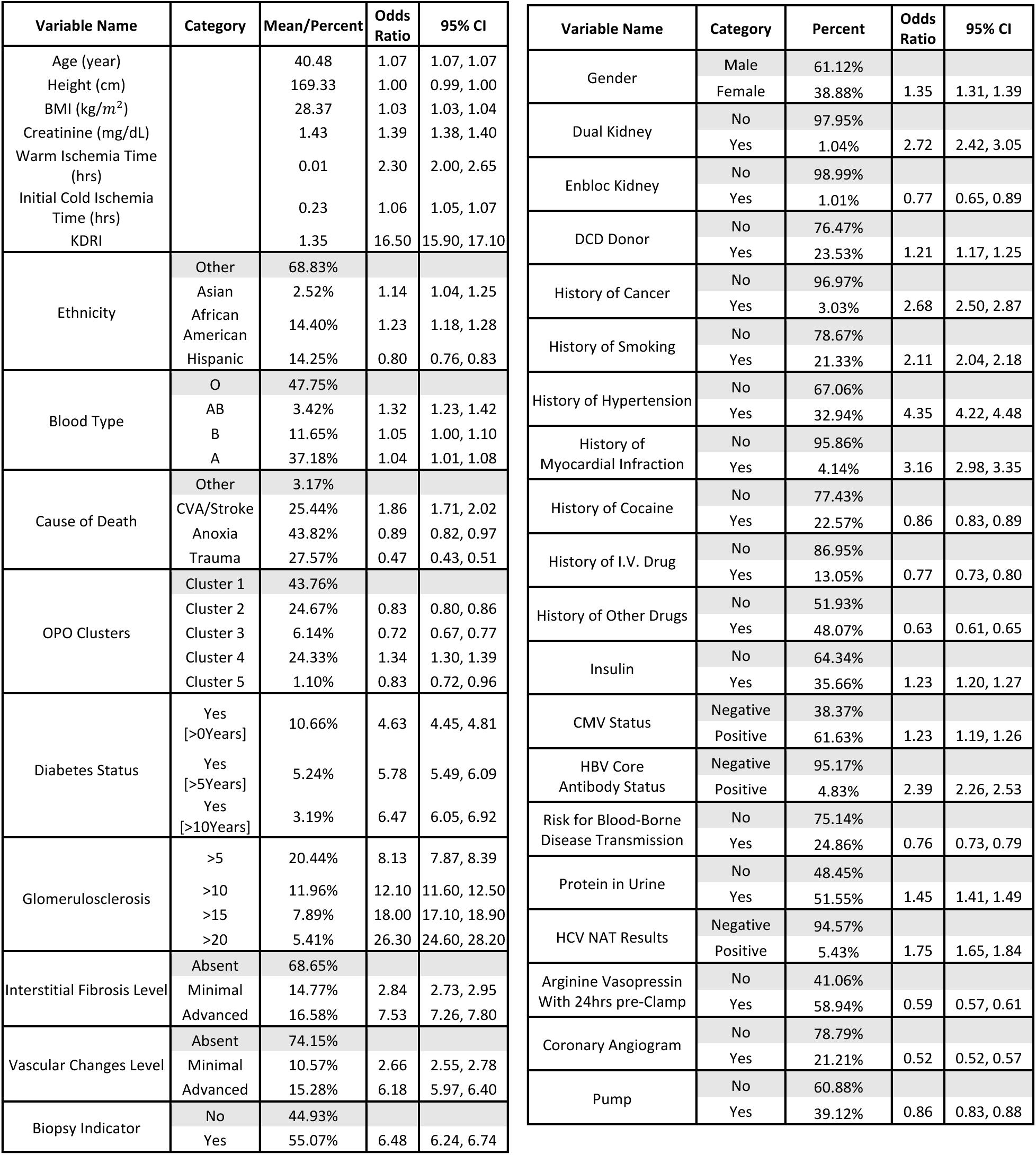
The results of the univariate analysis (N = 117 747). For continuous variables, the mean value is reported. For binary and categorical variables, the percentage of kidneys in each category is reported. The highlighted categories are used as the reference group when calculating the odds ratio for non-utilization.

### 2.2 Variable Creation

We created additional variables using some of the 36 variables included in our data. Specifically, we computed Cold Ischemia Time (CIT) and Warm Ischemia Time (WIT) for each kidney, since prolonged CIT and WIT adversely affect graft function,^23,24^ and transplant centers carefully evaluate these metrics when responding to kidney offers. For kidneys from Donation after Cardiac Death (DCD) donors, we calculated the WIT as the difference between agonal time and the clamp time. We calculated the CIT at the first match run creation (referred to as *CIT onset*) as the gap between the clamp time and the first match run creation time. CIT onset was set to zero if the clamp time is after the first match run creation.

En-bloc kidney transplantation is a procedure where two small kidneys from a donor weighing less than 18 kg are transplanted into one recipient.^25^ Dual (or 2-for-1) kidney transplantation involves transplanting both kidneys from a donor weighing at least 18 kg, which are individually less suitable for transplantation.^25^ To account for the disparities between the nonuse risk of en-bloc and dual kidneys, we created two indicator variables, namely *isEnbloc* and *isDual*.

Finally, we used the k-means method^26,27^ to cluster the OPOs based on two criteria: (i) the transplantation percentage of all kidneys they recovered, and (ii) the transplantation percentage of kidneys they recovered with KDPI greater than or equal to 85%. We created indicator variables for each cluster to study the effect of OPO characteristics on the utilization of kidneys.

### 2.3 Missing Data Imputation and Data Exclusion

Our data included variables with unspecified categories or categories that were deemed inconsequential to kidney disposition (e.g., the distinction between blood types A, A1, and A2). We consolidated those categories (see Table A1 in the online supplement). We further processed the data by replacing creatinine values above 20 with 20 for six kidneys; by computing missing BMI values using available donor weight and height; and by imputing missing WIT and CIT onset values using median observed values. After these steps, 1,406 kidneys (1.2%) with missing data were excluded from the analysis.

We excluded two kidneys with more than 24 hours WIT as outliers to avoid bias in our results. Furthermore, kidneys that were not used for reasons unrelated to their characteristics were excluded. In particular, we excluded kidneys with nonuse reasons “not as described” (0.05% of kidneys, 60 kidneys), or “recipient determined to be unsuitable in the operating room” (0.06% of kidneys, 76 kidneys). Lastly, categories with less than 20 observations were removed, as detailed in Table A1.

### 2.4 Model Development for Kidney Nonuse Risk Prediction

We employed two machine learning models for predicting nonuse risk: logistic regression (a parametric model) and random forest (a non-parametric model). For the logistic regression, non-linear effects of continuous variables were accounted for using linear splines (refer to Table A2 in the online supplement). We designed two classes of kidney nonuse risk prediction models:

#### Simplified Risk Models

These models use KDRI alongside a minimal set of variables, streamlining the risk assessment. For *random forest (RF)* models, initial training was done using all of the original variables (i.e., variables included in the UNOS data and created variables explained in Section 2.2) and KDRI. We then identified the top five, seven, and nine variables based on permutation importance. Models were then re-trained with these variables and KDRI. For *logistic regression (LR)* models, we included both the original and spline variables and compared the coefficients of normalized variables to identify the top ones.

#### Comprehensive Risk Models

Serving as a benchmark, these models excluded KDRI to avoid any implied, intrinsic biases or shortcomings of this metric. The *RF model* used all of the original variables, while the *LR model* used both the original and spline variables. The efficacy of simplified and comprehensive risk models was compared. Additionally, we evaluated the performance of our models against a benchmark nonuse risk prediction model that uses only KDRI, directly correlating it with nonuse risk, which we refer to as *KDRI alone*.

We performed five-fold stratified cross-validation and evaluated the performance of the proposed models using the receiver operating characteristic (ROC) and the precision-recall (PR) curves. The area under the ROC curve (AUROC) measures the model’s ability to distinguish between transplanted and unused kidneys. Precision and recall are critical metrics for prediction performance. Precision is equivalent to Positive Predictive Value (PPV); representing the ratio of correctly predicted unused kidneys to the total number of kidneys predicted as unused. Recall is equivalent to sensitivity; representing the ratio of correctly predicted unused kidneys to the actual number of unused kidneys. There is a trade-off between precision and recall; the proposed models can achieve higher recall at the expense of more false positives (i.e., transplanted kidneys predicted as unused) by lowering the prediction threshold. We plotted the PR curve for each model to visualize this trade-off and calculated the area under the PR curve (AUPRC).

### 2.5 Prediction Scenarios

We aim to predict kidney nonuse risk either before the beginning of the match run or during its early stages. While the majority of variables considered in our analysis are available pre-match-run, biopsy-related variables like glomerulosclerosis, interstitial fibrosis, and vascular change usually become available during the match run. The role of these variables in the kidney offer response decisions can be paramount,^4^ and more than half of the deceased donor kidneys undergo biopsy.^28^ We developed models with and without biopsy-related variables. In practice, decision-makers can readily utilize the models without biopsy-related variables before the beginning of the match run, and pivot to the ones with such variables once biopsy data become available.

### 2.6 Co-Design of Nonuse Risk Prediction Models

Risk prediction models developed by machine learning methods may prioritize certain variables over others based on statistical patterns in the data, which may not always align with clinical relevance. One reason for this is that the training data may not be totally representative of the population.^36^ To enhance the clinical relevance and interpretability of the proposed risk prediction models, we embarked on a co-design process with kidney transplant experts in our team. This collaborative effort entailed a systematic evaluation and potential substitution of variables. More precisely, transplant experts scrutinized the variables chosen by the machine learning algorithms and suggested replacing those deemed less relevant in kidney utilization with alternative variables. We assessed each recommended variable’s impact on prediction performance to ascertain the final set of model variables. The co-design approach fosters a deeper understanding of the clinical context, resulting in models that are not only accurate but also easily interpretable and actionable by healthcare providers.

## 3 Results

### 3.1 Study Population and Variable Analysis

Figure 2 outlines the data preparation process. Starting with 119,334 procured kidneys from 61 210 donors, we implemented four exclusion criteria removing 1,587 kidneys (1.33%). The final dataset consisted of 94,057 transplanted kidneys (79.9%) and 23,690 unused kidneys (20.1%). Figure 3 demonstrates the k-means clustering of the 58 OPOs into five groups based on the transplantation percentage of all kidneys and the transplantation percentage of hard-to-place (KDPI ≥ 85%) kidneys they recovered.

**Figure 3.**
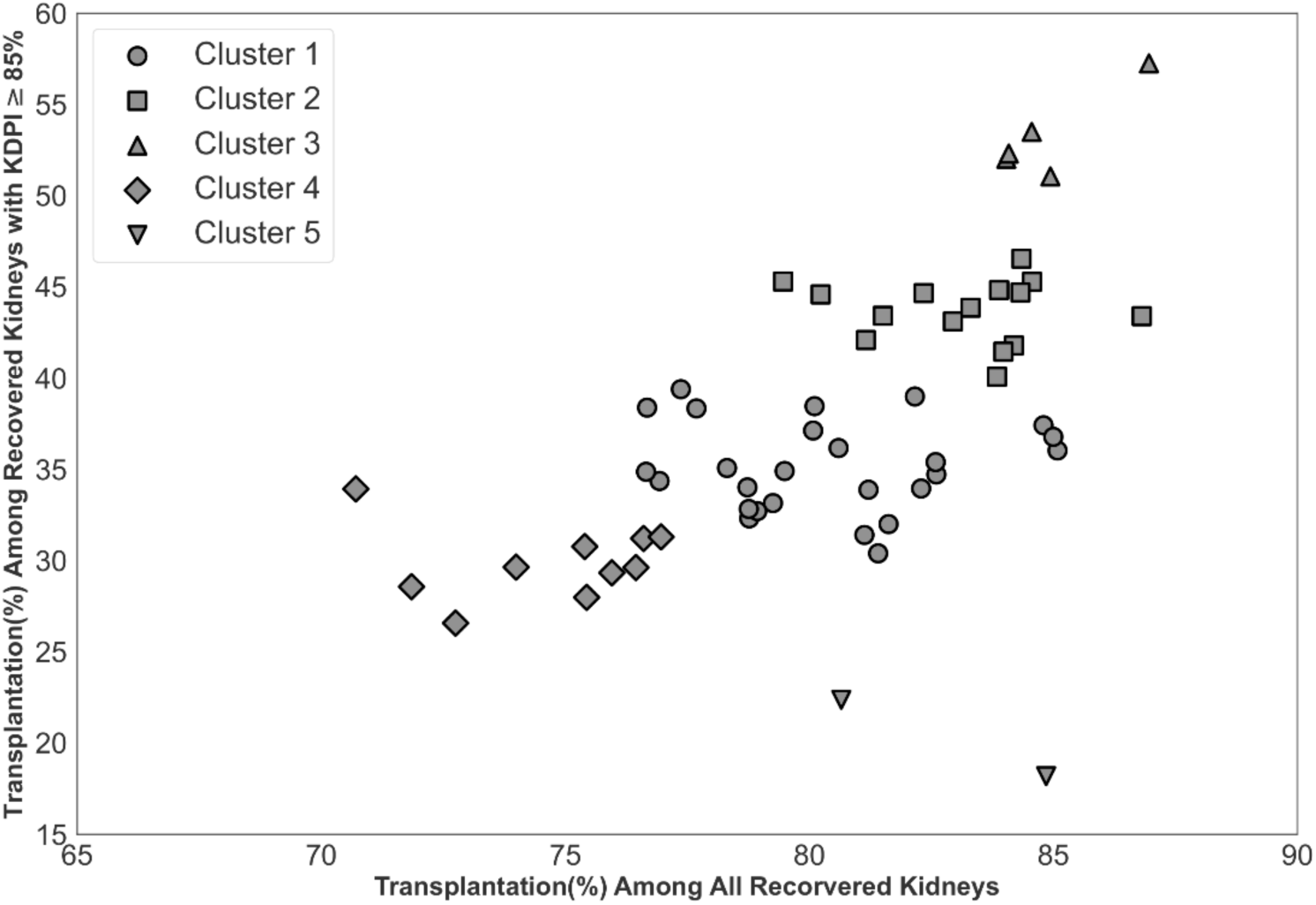
Impact of OPO centers on the disposition of hard-to-place kidneys. Categorizing OPO centers into five clusters to account for their impact.

Univariate analysis results (i.e., single factor analysis for nonuse) are displayed in Table 1. All variables exhibited statistically significant odds ratios for non-utilization. For instance, all else equal, with each hourly increment in WIT and CIT onset, we expect to see a 130% and 6% increase in the odds of nonuse as indicated by their odds ratios of 2.30 and 1.06, respectively.

### 3.2 Model Prediction Performance

Figure A1 presents the ROC and PR curves. The models that use KDRI and nine additional variables (*simplified risk models*) matched the performance of models trained on the entire variable set (*comprehensive risk models*). Figure 4 shows the performance of the models in the same plot for comparison. The RF model has the best performance, followed by the LR model-both exhibiting clearly superior performance compared to using KDRI alone. Figure A2 and Figure A3 show the impact of biopsy results on model performance: the performance of the LR model markedly improves whereas the performance of the RF model remains comparable. Table A3 reports the number of false positives avoided by the proposed models compared to using KDRI alone. For example, the RF model can precisely predict and potentially increase the transplantation likelihood of nearly 12,000 unused kidneys at a recall (sensitivity) level of 0.5. Concurrently, this model would prevent the misclassification of over 5,500 transplanted kidneys compared to using KDRI alone, mitigating thousands of needless interventions. The efficacy of the proposed prediction models becomes increasingly evident at higher recall levels.

**Figure 4.**
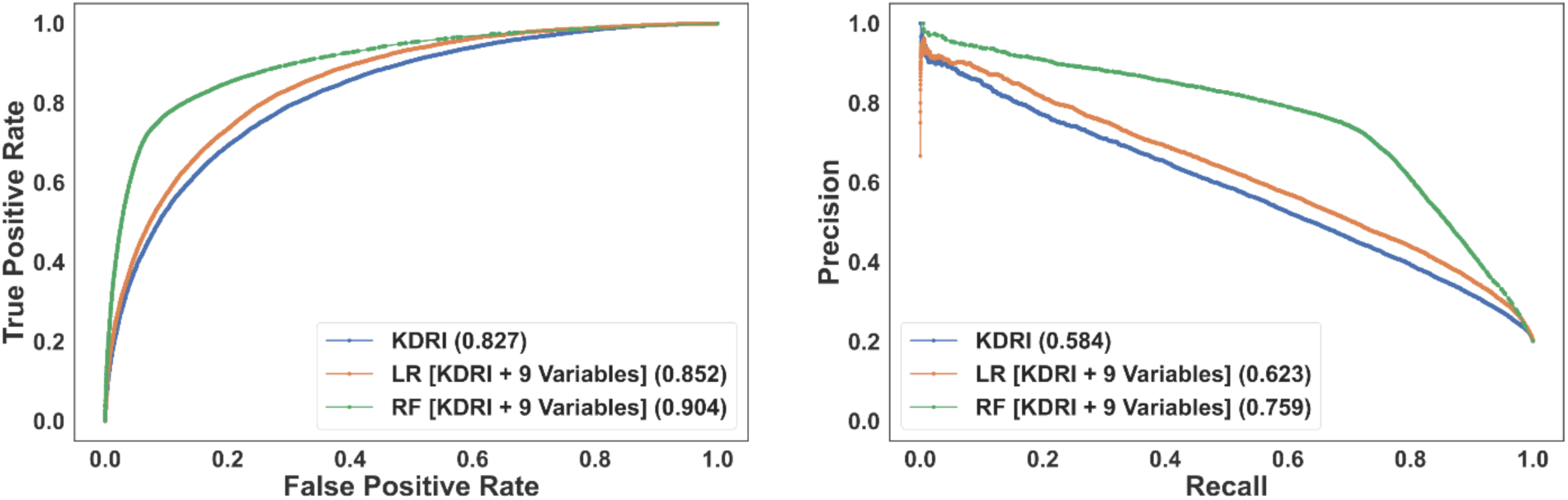
The ROC (left) and PR (right) curves for the simplified models incorporating KDRI and nine additional (non-biopsy-related) variables. The area under the curve of each model is reported in the legend.

### 3.3 Selected Variables

In the rest of this manuscript, we focus on the simplified RF model that includes KDRI and nine additional variables since its performance is clearly better than the simplified LR model with the same size. We denote the simplified RF model excluding the biopsy information as the *baseline model*. Table 2 displays the model variables in two prediction scenarios, including and excluding biopsy information. The variables in each column are reported in the order of their permutation importance. Creatinine, age, BMI, history of smoking, and height are chosen in both scenarios. In the model with biopsy information, glomerulosclerosis, interstitial fibrosis, and the biopsy indicator replaced the history of hypertension, coronary angiogram and DCD indicator variables.

**Table 2:** Variable selected by proposed simplified risk models when excluding and including biopsy-related variables. Variables that are commonly selected in both scenarios are given in bold.

|  | Baseline Model | Model With Biopsy Information |
| --- | --- | --- |
| 1 | **KDRI** | **KDRI** |
| 2 | **Creatinine** | **Creatinine** |
| 3 | **Age** | **Age** |
| 4 | History of Hypertension | Glomerulosclerosis |
| 5 | **Arginine** | Biopsy Performed |
| 6 | **BMI** | **BMI** |
| 7 | **History of Smoking** | Interstitial Fibrosis |
| 8 | DCD | **History of Smoking** |
| 9 | Coronary Angiogram | **Height** |
| 10 | **Height** | **Arginine** |

### 3.4 The Role of OPOs in the Disposition of Kidneys

The clustering analysis in Figure 3 demonstrated the differences between OPOs’ performance on kidney disposition. Furthermore, OPO cluster indicator variables had significant (unadjusted) odds ratios for nonuse in Table 1. To further assess the impact of OPO cluster variables, we computed their odds ratios for nonuse by adjusting for the nonuse risk predicted by the baseline model. Table A4 displays these adjusted odds ratios when using OPO cluster 1 as the reference group. Except for cluster 5, which has fewer observations than other clusters, all other clusters yielded significant risk-adjusted odds ratios for nonuse. The findings presented in Table A4 reveal, for example, that a kidney from an OPO in cluster 3 (cluster 4) is significantly less (more) likely to go unused compared to a similar kidney with the same predicted nonuse risk from an OPO in cluster 1.

### 3.5 Factors Increasing the Transplantation Likelihood of Kidneys at High Risk of Nonuse

In this section, we analyze the factors which increase the transplantation likelihood of hard-to-place kidneys. These factors can inform actionable interventions to improve the transplantation likelihood of such kidneys. We consider a kidney as hard-to-place if its risk prediction exceeds 0.75; when using this probability threshold, the number of identified kidneys that were not used (i.e., true positives) mirrors that were identified using the KDPI 85% benchmark. Specifically, of the 12 916 kidneys identified as hard-to-place, 10,845 (84%) were not used, while 2,071 (16%) were transplanted. We perform a univariate analysis among hard-to-place kidneys by adjusting for the nonuse risk. The results of this analysis are presented in Table 3, spotlighting factors that are associated with a higher transplantation likelihood of hard-to-place kidneys.

**Table 3:** Top significant factors that are associated with increased transplantation likelihood for hard-to-place kidneys. The odds ratio is adjusted for the nonuse risk.

| Variable Name | Adjusted Odds Ratio for Transplant | 95% CI | Percentage of transplanted hard-to-place kidneys when the variable value is YES | Percentage of transplanted hard-to-place kidneys when the variable value is NO/Baseline |
| --- | --- | --- | --- | --- |
| Enbloc Kidney | 5.88 | 1.76, 22.67 | 63% | 16% |
| Dual Kidney | 5.31 | 3.98, 7.07 | 52% | 16% |
| OPO Cluster3<br>(Baseline: OPO Cluster1) | 2.04 | 1.65, 2.51 | 28% | 16% |
| Pump | 2.01 | 1.82, 2.21 | 23% | 12% |
| OPO Cluster2<br>(Baseline: OPO Cluster1) | 1.13 | 1.00, 1.28 | 19% | 15% |

### 3.6 Outcomes of the Co-Design Experiment

Kidney transplantation experts in our team identified three variables of the baseline model, history of smoking, coronary angiogram, and height, as less relevant from a clinical perspective and recommended six potential alternative variables to replace them: OPO cluster, history of diabetes, cause of death, insulin, protein in urine, and pump. Similarly, for the model with biopsy information, eight potential variables were suggested (OPO clusters, history of diabetes, cause of death, insulin, protein in urine, pump, history of hypertension, and DCD indicator) to replace the two that were deemed less relevant (history of smoking and height). We evaluated 20 alternatives (six choose three) for the baseline model and 28 alternatives (eight choose two) for the model with biopsy information and selected the ones with the highest AUROC and AUPRC. Table 4 displays variables in these models. The modifications suggested by transplant experts did not compromise the predictive performance of our models; the AUC values remained comparable, and even increased slightly, for both the baseline model (0.905 versus 0.904 for AUROC and 0.767 versus 0.759 for AUPRC) and the model with biopsy information (0.905 versus 0.903 for AUROC and 0.771 versus 0.762 for AUPRC).

**Table 4:** Variables of the random forest nonuse risk prediction model after the co-design. Variables that are added to the model during the co-design process are in bold.

|  | Baseline Model | Model With Biopsy Information |
| --- | --- | --- |
| 1 | KDRI | KDRI |
| 2 | Creatinine | Creatinine |
| 3 | Age | Age |
| 4 | History of Hypertension | Glomerulosclerosis |
| 5 | Arginine | Biopsy Performed |
| 6 | BMI | BMI |
| 7 | DCD | Interstitial Fibrosis |
| 8 | **OPO Cluster** | Arginine |
| 9 | **Cause of Death** | **OPO Cluster** |
| 10 | **Pump** | **Pump** |

## 4 Discussion

Our study proposes kidney nonuse risk prediction models consisting of KDRI and nine additional variables. By achieving a balance between simplicity and performance, these models address a crucial gap in the organ allocation system—the need for easy-to-use yet accurate kidney nonuse risk prediction models. The proposed models can provide transparent and interpretable decision support to initiate interventions and manage allocation exceptions within the match-run system, and hence increase the transplantation likelihood of hard-to-place kidneys. Our simplified models significantly outperform using KDRI alone in predicting kidney nonuse risk, and exhibit performances on a par with substantially larger models with more variables.

If biopsy results are available, models with these variables can be used for nonuse risk prediction. However, the inclusion of biopsy information does not markedly enhance the performance of the RF models, suggesting that their predictive capability is robust regardless of the availability of such data. This challenges the conventional understanding in the literature that biopsy information is indispensable for nonuse risk prediction.^4,29^ The impact of biopsy results on the utilization of kidneys does, however, manifest clearly in the LR models. That is, the significance of biopsy results for nonuse risk prediction varies across modeling approaches.

In addition to KDRI, terminal creatinine level, age, BMI, and use of arginine vasopressin within 24 hours pre-cross clamp are significant predictors of kidney nonuse risk. If biopsy is performed, our models also utilize variables such as interstitial fibrosis and glomerulosclerosis, aligning well with previous studies that emphasized the role of glomerulosclerosis in kidney non-utilization.^30,31^ If the biopsy results are not available, then the models utilize variables such as the DCD indicator and history of hypertension. It is worth noting that some of the variables used in our models, such as age, creatinine, and history of hypertension are also considered in KDRI calculation. The predictive performance gap between the proposed models and using KDRI alone emphasizes the importance of recalibrating KDRI for nonuse risk prediction.

Our prediction results reveal the significance of OPO-related factors in the utilization of hard-to-place kidneys. We identify a cluster of OPOs that demonstrate exemplary performance in placing such kidneys. The inclusion of OPO-related factors in our risk prediction models is not just a technical innovation but a call to action for the transplantation community to analyze and disseminate the successful strategies of high-performing OPOs, thereby elevating overall practice standards and to encourage other OPOs to adopt similar, effective approaches in organ recovery and allocation. For example, the literature documents major disparities in making out-of-sequence kidney offers to accelerate the placement of hard-to-place kidneys.^32^ Our models can help mitigate such disparities by providing guidance to OPOs for identifying hard-to-place kidneys that can be intervened for better utilization and for standardizing interventions to enhance transparency and equitability.

The results of the model co-design approach confirm that data-driven machine learning methods and clinical expertise are not mutually exclusive but complementary. By incorporating the insights of transplant experts into the model development process, we have created models that not only have high prediction performance but also align well with real-world clinical judgments, enhancing the medical relevance of our results. In particular, three variables that are deemed less relevant to kidney utilization, the history of smoking, coronary angiogram, and height (when considered in addition to BMI), are replaced with OPO cluster, cause of death, and pump indicator variables through the co-design process.

After identifying hard-to-place kidneys in our data using the proposed nonuse risk prediction models, we explored their characteristics that are associated with an increased transplantation likelihood within the current allocation system. These characteristics can inform the development of *operational* and *system-level* interventions. Operational interventions that are identified in our analysis include pumping kidneys, which can help maintain graft function,^33^ and presenting dual offers, which can increase the chances of acceptance.^34^ System-level interventions require strategic changes at the policy or organizational level and often involve a longer-term approach. Such interventions identified in our analysis include identifying and promoting the best organ recovery and allocation practices across OPOs or the integration of an effective nonuse risk prediction framework into the national allocation system.

Nevertheless, our study is not without limitations. Our dataset, spanning from 2016 to 2021, does not completely capture the potential impacts of recent policy changes post-March 2021. Additionally, the observed decline in nonuse rates for hepatitis C-positive donor kidneys, attributable to treatment advances,^35^ suggests that our models might benefit from updated datasets to reflect current trends. Moreover, while our models aim to enhance the utilization of kidneys at high risk of nonuse, it is essential to balance this goal with the imperative to avoid adverse outcomes for recipients. The integration of lower-quality kidneys into the transplant pool necessitates careful consideration to prevent complications and ensure recipient welfare. Ongoing collaborations with medical decision-makers, organ transplant researchers, and ethics committees will be vital in navigating these challenges.

In summary, our work presents interpretable models that effectively predict the nonuse risk of deceased donor kidneys, thereby paving the way for timely data-driven interventions that can alleviate the alarming rates of kidney nonuse. Although these models consist of a small number of variables including KDRI along with factors selected by machine learning models and transplant experts, they perform comparably to more complex risk prediction models. The integration of these simplified models into organ procurement and allocation processes can enhance organ utilization and mitigate disparities in access to transplantation. Future research is warranted to validate these findings with newer datasets, evaluate the impact of recent policy changes, and assess the real-world implementation of nonuse risk prediction models in the actual organ allocation systems.

## Data Availability

This study is based on OPTN data as of October 1, 2021. We comply with all the provisions specified in the "Data User Agreement" provided by the OPTN. In short, the authors can neither use nor permit others to use the Data in any way other than for statistical reporting and analysis. We can neither release nor permit others to release the Data to any person (including media and subcontractors) except with the written approval of UNOS.
The data reported here have been supplied by the United Network for Organ Sharing as the contractor for the Organ Procurement and Transplantation Network. The interpretation and reporting of these data are the responsibility of the author(s) and in no way should be seen as an official policy of or interpretation by the OPTN or the U.S. Government.

