## Appendices for "Improving Deceased Donor Kidney Utilization: Predicting Risk of Nonuse with Interpretable Models"

***Table A1:*** *The original and final categories of the variables included in the analysis.*

| Variable Name | Original Category | Count | Category |
|---|---|---|---|
| Ethnicity | White | 80031 | Other |
| | Multiracial | 1032 | |
| | Amer Ind/ Alaska Native | 739 | |
| | Native Hawaiian/ other Pacific Islander | 365 | |
| | Asian | 17218 | Asian |
| | African American | 16970 | African American |
| | Hispanic | 2979 | Hispanic |
| Blood Type | O | 56959 | O |
| | A1 | 21076 | A |
| | A | 18797 | |
| | A2 | 4488 | |
| | B | 13925 | B |
| | AB | 1937 | AB |
| | A1B | 1434 | |
| | A2B | 718 | |
| Cause of Death | Anoxia | 52334 | Anoxia |
| | Trauma | 32836 | Trauma |
| | CVA/Stroke | 30337 | CVA/Stroke |
| | Other | 3426 | Other |
| | Tumor | 401 | |
| Diabetes Status | No | 105387 | No |
| | Unknown | 1183 | |
| | Yes [0-5Years] | 4914 | Yes [>0Years] Yes [>5Years] Yes [>10Years] |
| | Yes [6-10Years] | 2448 | |
| | Yes [>10Years] | 3819 | |
| | Yes [Unknown] | 1583 | |

| Variable Name | Original Category | Count | Category |
|---|---|---|---|
| History of Hypertension | No | 78683 | No |
| | Unknown | 1286 | |
| | Yes | 39365 | Yes |
| History of Cancer | No | 114554 | No |
| | Unknown | 1172 | |
| | Yes | 3608 | Yes |
| History of Myocardial Infraction | No | 111544 | No |
| | Unknown | 1617 | |
| | NaN | 1279 | |
| | Yes | 4894 | Yes |
| History of Cigarette | No | 91202 | No |
| | Unknown | 2650 | |
| | Yes | 25480 | Yes |
| | NaN | 2 | Remove |
| History of Cocaine | No | 89130 | No |
| | Unknown | 2313 | |
| | NaN | 1262 | |
| | Yes | 26629 | Yes |
| History of I.V. Drug | No | 101805 | No |
| | Unknown | 1948 | |
| | Yes | 15575 | Yes |
| | NaN | 6 | Remove |
| History of Other Drug | No | 59661 | No |
| | Unknown | 1681 | |
| | NaN | 1262 | |
| | Yes | 56730 | Yes |
| HCV NAT Results | Negative | 112820 | Negative |
| | Positive | 6491 | Positive |
| | Not Done | 12 | Remove |
| | Indeterminant | 9 | Remove |
| | Unknown | 2 | Remove |

| Variable Name | Original Category | Count | Category |
|---|---|---|---|
| Insulin | No | 62527 | No |
| | NaN | 14365 | |
| | Yes | 42442 | Yes |
| HBV Core Antibody Status | Negative | 113430 | Negative |
| | Not Done | 124 | |
| | Positive | 5776 | Positive |
| | Indeterminant | 4 | Remove |
| Arginine Vasopressin With 24hrs pre-Clamp | Yes | 69561 | Yes |
| | No | 48463 | No |
| | NaN | 1274 | |
| | Unknown | 36 | |
| Coronary Angiogram | No | 93051 | No |
| | NaN | 1275 | |
| | Yes | 25008 | Yes |
| Protein in Urine | Yes | 60886 | Yes |
| | No | 56450 | No |
| | NaN | 1262 | |
| | Unknown | 736 | |
| Risk for Blood-Borne Disease Transmission | No | 89756 | No |
| | Yes | 29572 | Yes |
| | Unknown | 6 | Remove |
| CMV Status | Positive | 72966 | Positive |
| | Indeterminant | 490 | |
| | Negative | 45794 | Negative |
| | Not Done | 82 | |
| | Unknown | 2 | Remove |

| Variable Name | Original Category | Count | Category |
|---|---|---|---|
| Kidney Biopsy | Yes | 65606 | Yes |
| | No | 50910 | No |
| | NaN | 2818 | |
| Kidney Percentage of Glomerulo-sclerosis | Unknown | 51818 | 0-5 |
| | 0-5 | 40602 | |
| | Not Reported | 2468 | |
| | Indeterminate | 280 | |
| | 20+ | 6409 | >20 |
| | 6-10 | 10028 | >5 |
| | 11-15 | 4800 | >10 |
| | 16-20 | 2929 | >15 |
| Kidney Interstitial Fibrosis | Unknown | 51805 | Absent |
| | Absent | 27534 | |
| | Not Reported | 2468 | |
| | Unknown | 482 | |
| | Minimal | 17447 | Minimal |
| | Mild | 15129 | Advanced |
| | Mild-moderate | 3921 | |
| | Severe | 548 | |
| Kidney Vascular Changes | Unknown | 51807 | Absent |
| | Absent | 32192 | |
| | Not Reported | 2468 | |
| | Unknown | 2312 | |
| | Minimal | 12489 | Minimal |
| | Mild | 12263 | Advanced |
| | Mild-moderate | 4793 | |
| | Severe | 1010 | |

***Table A2:*** *Linear splines considered in the logistic regression models.*

| Variable Name | Linear Spline (LS) | |
|---|---|---|
| Age | > 55 | right LS |
| | > 65 | right LS |
| Height | < 160 | left LS |
| BMI | > 30 | right LS |
| Creatinine | > 1 | right LS |
| | > 2 | right LS |
| WIT | > 0.5 | right LS |
| | > 1 | right LS |
| Initial CIT | > 5 | right LS |
| | > 10 | right LS |

*Notes. Example spline calculation: Consider age and height, which are continuous variables. Assume that we want to add the right linear spline variable at age 55. For the donor $i$ with age $A_i$, the right linear spline variable takes the positive part of $(A_i - 55)$. Similarly, if we want to add the left linear spine variable at height 160, the variable takes the positive part of $(160 - H_i)$ for $H_i$ denotes the donor height.*

***Table A3:*** *The number of false positives avoided by proposed prediction models without biopsy-related variables compared to using only the KDRI. Recall represents the faction of all unused kidneys that are correctly classified.*

| Recall | RF (KDRI + 9 Variables) | LR (KDRI + 9 Variables) |
|---|---|---|
| 0.5 | 5739 | 1395 |
| 0.6 | 9078 | 2202 |
| 0.7 | 13694 | 3151 |
| 0.8 | 17136 | 5033 |

***Table A4:*** *Adjusted odds ratio for cluster indicators. The odds ratio is adjusted for the predicted risk.*

| Variable Name | Adjusted Odds Ratio for Nonuse | 95% CI |
|---|---|---|
| OPO Cluster2 | 0.86 | 0.81, 0.90 |
| OPO Cluster3 | 0.81 | 0.74, 0.89 |
| OPO Cluster4 | 1.13 | 1.07, 1.18 |
| OPO Cluster5 | 1.05 | 0.85, 1.28 |

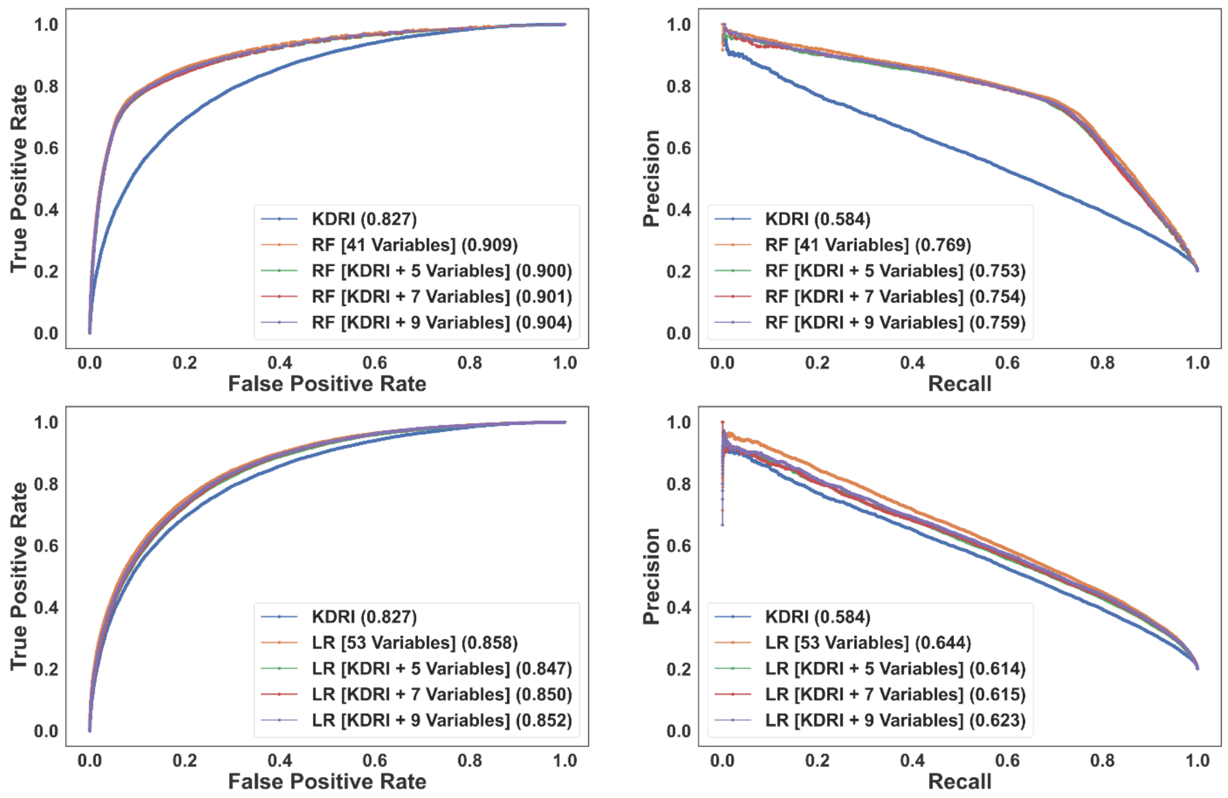


***Figure A1*** *The ROC (left) and PR curves (right) for random forest and logistic regression models without biopsy-related variables. The area under the curve of each model is reported in the legend.*

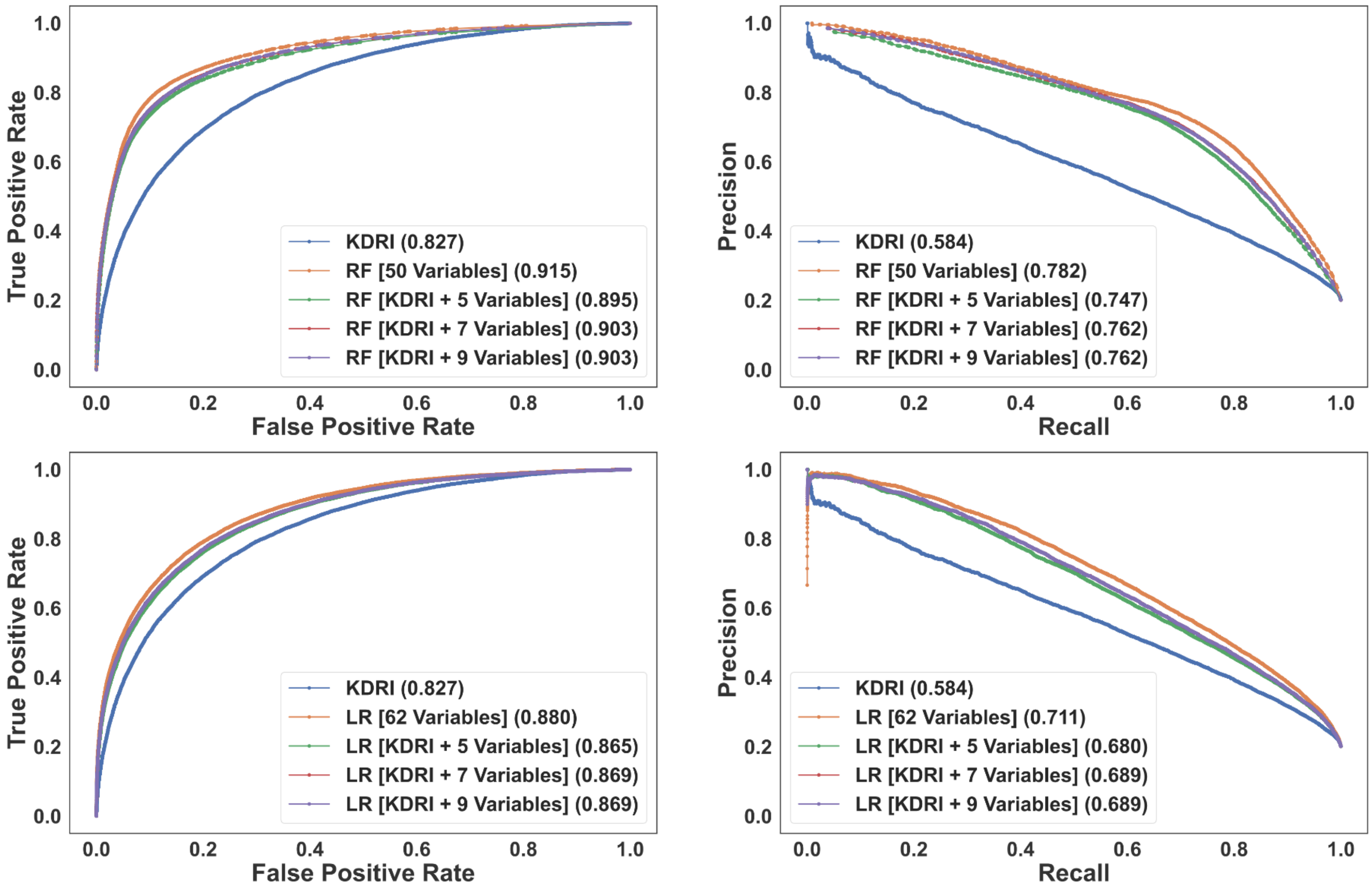


***Figure A1:*** *The ROC (left) and PR curves (right) for random forests and logistic regression models when biopsy information is available. The area under the curve of each model is reported in the legend.*

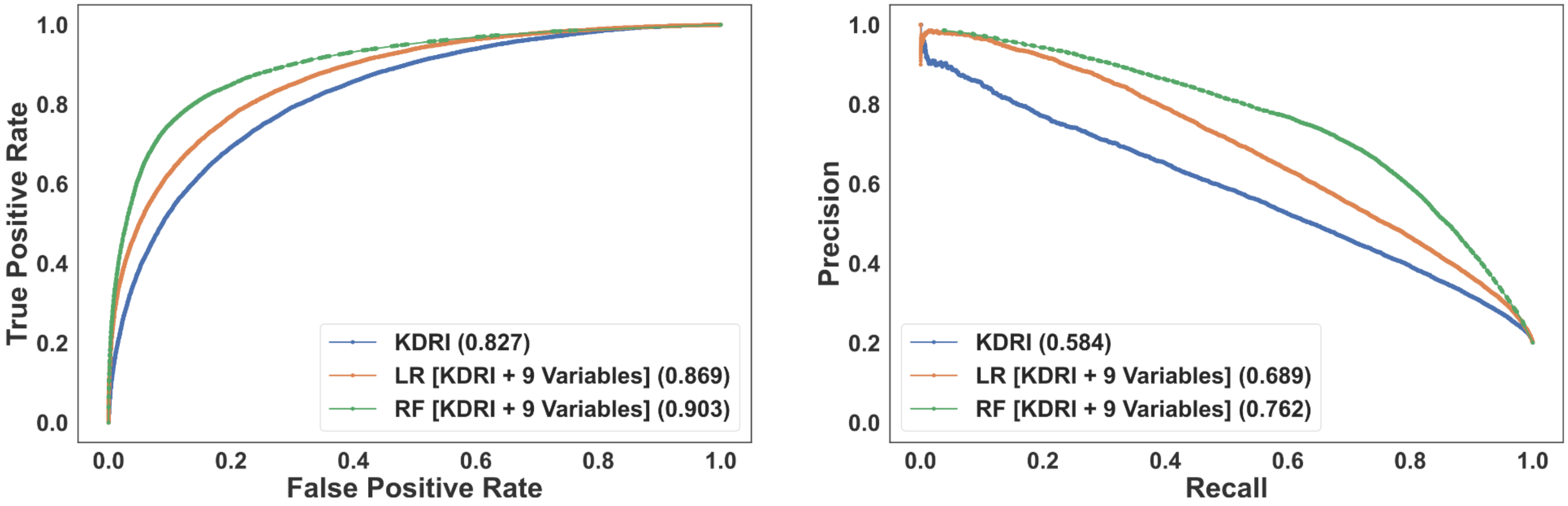


***Figure A2:*** *The ROC (left) and PR (right) curves for the simplified models incorporating KDRI and nine additional variables when biopsy information is available. The area under the curve of each model is reported in the legend.*
